# Complement-dependent mpox virus-neutralizing antibodies in infected and vaccinated individuals

**DOI:** 10.1101/2023.04.14.23288385

**Authors:** Mathieu Hubert, Florence Guivel-Benhassine, Timothée Bruel, Françoise Porrot, Delphine Planas, Jessica Vanhomwegen, Aurélie Wiedemann, Sonia Burrel, Stéphane Marot, Romain Palich, Gentiane Monsel, Harouna Diombera, Sébastien Gallien, Jose Luis Lopez-Zaragoza, William Vindrios, Fabien Taieb, Sandrine Fernandes-Pellerin, Maurine Delhaye, Hélène Laude, Laurence Arowas, Marie-Noelle Ungeheuer, Laurent Hocqueloux, Valérie Pourcher, Thierry Prazuck, Anne-Geneviève Marcelin, Jean-Daniel Lelièvre, Christophe Batéjat, Yves Lévy, Jean-Claude Manuguerra, Olivier Schwartz

## Abstract

Mpox virus (MPXV) caused a multi-country outbreak in non-endemic areas in 2022. The Modified Vaccinia Ankara (MVA)-based vaccine was used as prophylaxis, but its effectiveness remains poorly characterized. Here, we developed two assays for quantification of neutralizing antibodies (NAbs), using MVA-GFP or a recently isolated MPXV. We measured NAb levels in 470 sera from control, MPXV-infected or MVA-vaccinated individuals. Various levels of MVA NAbs were detected after infection, historic smallpox or MVA vaccination. MPXV was barely sensitive to neutralization. Addition of complement enhanced detection of responsive individuals and NAb levels. Anti-MVA and -MPXV NAbs were observed in 94% and 82% of infected individuals, respectively, and 92% and 56% of MVA vaccinees, respectively. NAb titers were higher in individuals born before 1980, highlighting the impact of historic smallpox vaccination on humoral immunity. Altogether, our results indicate that MPXV neutralization is complement-dependent and help uncover the mechanisms underlying vaccine effectiveness.

**SUMMARY:** In 2022, mpox virus (MPXV) caused an unprecedented pandemic outbreak in non-endemic areas. The efficacy of currently available third generation MVA-based vaccines and the nature of the humoral response generated after MPXV infection remain poorly characterized. We established cell-based assays to measure neutralizing antibodies (NAbs) targeting MVA or MPXV. We analyzed 470 sera and detected robust levels of MVA NAbs after infection, historic smallpox vaccination or administration of MVA-based vaccines. Efficient MPXV neutralization required addition of complement. High NAb titers were measured in ancient smallpox-vaccinated MPXV-infected patients, suggesting a potential cross-protection mediated by hybrid immunity.

## INTRODUCTION

Cessation of massive vaccination in 1980 after smallpox eradication increased the risk of an *Orthopoxvirus* (OPXV) emergence in humans. Monkeypox virus, now termed mpox virus (MPXV), is a zoonotic virus that circulates in various animal species, mainly rodents, living in African rainforests. MPXV is endemic and in expansion in human populations from Central and West Africa^1^. Two clades of MPXV were initially described. Clade 1 (ex-Central Africa clade) is associated to most of African outbreaks and causes about 11% lethality. Clade 2 (ex-West Africa clade) has a lower fatality rate (1-6%) and is the most exported clade^2^. The introduction of MPXV into non-endemic areas was successively reported in 2003 (USA^3^), 2018 (UK^4^, Israel^5^, Singapore^6^) and 2021 (UK^4^, USA^7,8^). In May 2022, a genotypically distinct MPXV (clade 2b) was first detected in Europe and spread all over the world^2,9^. About 85,000 cases were reported in 110 countries, prompting the World Health Organization to claim a public health emergency of international concern. Identification of the high-risk population, Men having Sex with Men (MSM), bisexuals and their sexual networks, allowed adapted communication and vaccination campaigns, that were associated with the end of the outbreak in late 2022. People living with HIV accounted for 38-50% of the cases^10^. The case fatality rate was low (0.1%) but most of severe and fatal cases occurred in individuals with undiagnosed advanced HIV infection^10–12^. About 97% of cases reported during this recent outbreak were young men (median age 34), thus not previously vaccinated against smallpox. A surveillance study conducted on 760 mpox cases between 2005-2007 in Democratic Republic of Congo (DRC) showed that 3.8 % of the infected individuals had previous smallpox vaccination compared with 26.4% of the overall population^13^. Previously vaccinated individuals had a 5.2-fold lower risk of MPXV infection than unvaccinated counterparts. A 20-fold increase in mpox incidence was observed in DRC between 1980s and 2007, that may be linked to decreasing herd immunity^13^. However, 21% of infected individuals reported during the 2003 surge in USA were previously vaccinated^14^, indicating that childhood vaccination does not fully protect against MPXV. The historic smallpox vaccination was performed with first- and second-generation vaccinia virus (VACV)-based vaccines. A cross-protection of about 80% against mpox disease was reported^13,15^. The protection slowly wanes over time, leading to pauci-symptomatic infection^14,16,17^. The memory B cell response generated by smallpox vaccine is however long-lasting^18,19^. Depending on studies, up to 90% of individuals vaccinated 25–75 years ago maintain substantial humoral or cellular immunity against VACV^20–22^. Current third-generation vaccines are based on an attenuated Modified Vaccinia virus Ankara (MVA). The attenuated virus contains 6 major deletions compared to the ancestral VACV, representing a 10 % loss of its genome^23,24^. MVA was initially developed to serve as a safer vaccine during the last years of the WHO smallpox eradication campaign. A**n** MVA-based vaccine produced by Bavarian Nordic (MVA-BN, commercialized under the names of IMVANEX or JYNNEOS) was FDA-approved for extended use against MPXV on the basis of challenge experiments in non-human primates (NHP)^25–27^. Only **a** few studies investigated MVA-based vaccine effectiveness in humans. In Israel, a short-term study estimated that one MVA dose provided as pre-exposure prophylaxis was associated with 79% reduction in infection rates^28^. Another study conducted in USA during the 2022 outbreak monitored more than 5,000 mpox cases and showed a 14-fold increase in infections of unvaccinated individuals compared to JYNNEOS recipients (single dose regimen)^29^. The epidemiological studies estimating vaccine effectiveness are based on mpox incidence, but the underlying mechanisms remain poorly characterized. In non-human primates (NHP) having received a second generation vaccine, B-cell depletion experiments demonstrated that protection against severe mpox disease was mediated by neutralizing antibodies (NAbs)^30^. Depletion of CD4^+^ and/or CD8^+^ T cells did not impact vaccine efficacy. In a late model of infection in which NHP**s** were infected with MPXV 3 years after vaccination, no correlation was observed between pre-challenge VACV-specific cellular immunity and peak viremia^31^. In humans, acute MPXV infection triggers a rapid expansion of effector memory CD4^+^ and CD8^+^ T cells and an intensive Th1-biased response with exacerbated levels of inflammatory mediators (IL-1β, IL-6, IL-8, TNF)^32^. Whether MVA-based vaccines trigger similar cellular responses remains to be fully understood. To explore the humoral responses in vaccinated or infected individuals, quantification of the levels of circulating antibodies is often assessed by home-made ELISA or sero-neutralization assays directed against VACV, for reglementary reasons. Some assays are also performed with a recombinant virus carrying a GFP reporter gene (MVA-GFP) allowing easy monitoring of infection. In a recent study, low levels of MPXV-neutralizing antibodies were detected after MVA vaccination in healthy individuals^33,34^, but neutralization assays were performed in the absence of complement. The complement was previously shown to be required for VACV neutralization by monoclonal antibodies and plasma from VACV-infected mice or vaccinated humans, but its role against MPXV is less characterized^35–42^.

Here, we developed two assays for quantification of NAbs against MVA and the new clade of MPXV, in presence or absence of complement. We compared NAbs levels in sera from healthy donors having either received or not received the historical smallpox vaccine, MPXV-infected patients, and MVA-vaccinated individuals. We highlight the role of the complement for efficient MPXV neutralization.

## RESULTS

### Cohorts design

#### Uninfected individuals (CORSER-4 cohort)

The interventional cohort clinical study (CORSER-4) was initiated in 2021, with the primary objective to study humoral responses after SARS-CoV-2 infection or vaccination in healthy individuals. A secondary objective was to analyze the basal humoral response against other viruses, including MPXV, for which no known risk of viral exposure was reported in the cohort. We randomly selected 88 individuals in two categories of age. Thirty-four individuals were born after 1980, the cessation of smallpox vaccination, and 54 were born before this year and thus most likely vaccinated against smallpox during their childhood (Table 1).

#### MPXV-infected individuals

A total of 66 plasma or sera was collected from 48 MPXV-infected patients diagnosed in two hospitals: 57 sera or plasma from 39 patients at Hôpital Pitié-Salpêtrière (Paris) and 9 sera from 9 patients at Hôpital Henri Mondor (Créteil). Most of patients (69%) were sampled at a single time-point (3 to 33 days after onset of the symptoms, DOS) while 31% were sampled 2 to 4 times. The main characteristics of the patients are depicted in Table 2. Plasmatic MPXV genome was detectable by PCR from 2 to 25 DOS (median DOS:5 days) (Extended Data Fig. 1a).

Some patients cleared the MPXV genome from the bloodstream as soon as 3 DOS and PCR-negative plasma samples were collected from 3 to 80 DOS (median DOS: 20 days) (Extended Data Fig. 1a). We observed that 75% and 91% of infected individuals cleared plasmatic viral DNA 3 or 4 weeks after illness onset, respectively (Extended Data Fig. 1b), consistently with previous observations^43^. The clinical or molecular diagnosis of MPXV infection was performed 2 to 15 DOS (Extended Data Fig. 1c). This timeframe may explain MPXV transmission dynamics in populations at risk.

#### IMVANEX recipients

Sera from IMVANEX vaccinees were collected at the Centre Hospitalier Universitaire d’Orléans, and at Hôpital Henri Mondor. The characteristics of the patients are indicated in Table 3. We analyzed 147 sera from 86 individuals that were not infected by MPXV at the time of inclusion. Most of older individuals (born before 1980) received a single dose of IMVANEX except 4 who received a second dose. The 32 younger individuals (born after 1980) were not previously vaccinated and received two doses within 28 to 42 days of each other.

#### MVA-HIV recipients (VRI01 clinical trial)

We analyzed 169 sera from 67 participants of a clinical trial performed in 2014-2015. Participants received two doses of an MVA-HIV vaccine candidate separated by 8 weeks^45^. In total, 66 sera were collected at day 0 to measure pre-existing or basal neutralization levels, 42 sera were collected 2 weeks after the 1^st^ dose and 61 after the 2^nd^ dose (Table 4).

### Development of MVA and MPXV neutralization assays

We set up two assays to measure the neutralizing activity of sera or plasma against MVA and MPXV. Samples were heated 30 min at 56°C to inactivate the complement and interferons that may be potentially present.

The first assay was based on the use of a MVA virus expressing GFP upon infection (MVA-GFP)^47^, allowing a rapid quantification of viral infection. We selected Vero E6 cells as target cells because they are naturally sensitive to MVA and widely used in neutralization assays. The principle of the neutralization assay is outlined in Extended Data Fig. 2A. MVA-GFP particles were exposed for 2h at 37°C to serial dilutions of the sera, with or without 10% guinea pig serum as a source of complement (GPC)^35,42,49^. The mixture was then added onto Vero E6 cells. After 20h, the area of GFP^+^ cells was scored with an automated microscope. We first determined the optimal viral input by exposing Vero E6 cells to increasing doses of MVA-GFP. The number of GFP^+^ cells correlated with the viral inoculum (Extended Data Fig. 2B). We show two examples of a non-neutralizing serum and a neutralizing serum collected from one MPXV-infected patient at days 2 and 56 DOS, respectively (Extended Data Fig. 2C). We then calculated the effective dose of serum required to inhibit 50% of infection (ED50) and demonstrated an increase in ED50 overtime (Extended Data Fig. 2C). Sera from uninfected individuals did not neutralize MVA (see below). Neutralization across the different viral inocula was similar, indicating that variations in the number of infected cells, at least in this selected range of infection, did not impact the calculation of ED50 (Extended Data Fig. 2D). Neutralization was about 2.5-fold more efficient in the presence of complement, independently of the viral inoculum (Extended Data Fig. 2E). Heat-inactivated guinea pig sera did not enhance neutralization titers of the tested sera (not shown).

The second assay used an authentic clinical MPXV strain isolated on Vero E6 cells from a typical lesion of a French patient infected in June 2022 (genbank accession number OQ249661). The virus was cytopathic and formed plaques in Vero E6 cells, with titers reaching 5.10^7^ plaque forming units (PFU)/mL. For the neutralization assay, we tested different cell lines and observed that the U2OS human osteosarcoma-derived cell line was particularly sensitive to MPXV infection. The flat shape of U2OS cells facilitated automated imaging^50^. Clusters of infected cells, revealed after immunostaining with a rabbit polyclonal anti-VACV antibody, were readily detectable two days after infection. The MPXV neutralization assay is thus roughly similar to the MVA assay, except that target cells and incubation times are different, as outlined in Extended Data Fig. 3A. In the absence of serum, the number of infected cells correlated with the viral inoculum (Extended Data Fig. 3B). We selected a non-saturating inoculum for neutralizing experiments. Infected cells formed foci of different size, reflecting the known direct cell-cell transmission of poxvirus^51^. Contrary to the abortive single-round MVA infection in mammalian cells, MPXV infection leads to continuous production of infectious particles over time, introducing a variability in the number of infected cells and size of the foci. The automated scoring of infection based on the surface of positive cells was thus unsuitable to accurately calculate a neutralization activity. We thus defined a binary MPXV neutralization index for each serum dilution, by considering the “no serum” infection signal as negative (non-neutralizing), whereas wells in which the infection signal was reduced were considered as positive (neutralizing). The neutralization titer was thus calculated as the highest sample dilution in which neutralization was detected. Examples of non-neutralizing and neutralizing sera are presented in Extended Data Fig. 3C. With both MVA and MPXV assays, we arbitrarily defined a positive threshold (Limit of Detection or LOD) for samples displaying a neutralizing activity at 1/40 dilution, two-fold above the first dilution of sera tested. We selected this cut-off to avoid unspecific neutralization results that may be caused by high concentrations of human sera. The same threshold was used for experiments performed with or without complement. The proportion of neutralizers was then evaluated by calculating the percentage of individuals exhibiting a neutralizing activity > LOD.

We used the two assays to measure the neutralizing activity of 470 sera from healthy controls and from different categories of MPXV-infected or MVA-vaccinated individuals. Neutralization titers were calculated with or without complement supplementation.

### Basal neutralizing activity of samples from uninfected or unvaccinated individuals

We first assessed the basal neutralization levels of sera from uninfected donors (Fig. 1). We took advantage of the different cohorts to analyze a total of 195 sera. We combined 88 sera from the CORSER-4 cohort of uninfected donors, 41 sera of IMVANEX vaccinees collected before vaccination, and 66 sera of MVA-HIV vaccinated individuals collected before administration of the candidate vaccine. We separated the donors by age: 95 donors born before 1980 were likely vaccinated in their childhood and were designed as “older” participants, whereas 100 donors born after 1980, or “younger” participants, were considered as unvaccinated. Fig. 1A displays the neutralization titers for each individual and Fig. 1B the percentage of individuals with a detectable neutralizing activity. In the absence of complement, we did not detect anti-MVA NAbs in 99% (99/100) of younger donors. Low levels of NAbs (mean ED50: 127) were measured in 12% (11/95) of older donors, suggesting a residual humoral response to historical smallpox vaccination. Addition of 10% guinea pig serum as a source of complement enhanced neutralization titers to a mean ED50 of 273 in 3% (3/100) and a mean of 494 in 44% (42/96) of younger and older individuals, respectively. Thus, the percentage of MVA neutralizers among older individuals was approximatively 3-fold higher in the presence of complement. With MPXV, we did not detect any neutralization with these sera in the absence of complement. Addition of complement enhanced neutralization titers to a mean of 51 in 7% (7/100) of younger participants and a mean of 70 in 27% (26/95) of older participants. In 5/7 younger individuals that neutralized MPXV, titers remained low (equal to 40), and may thus represent a non-specific neutralization.

**Fig. 1.**
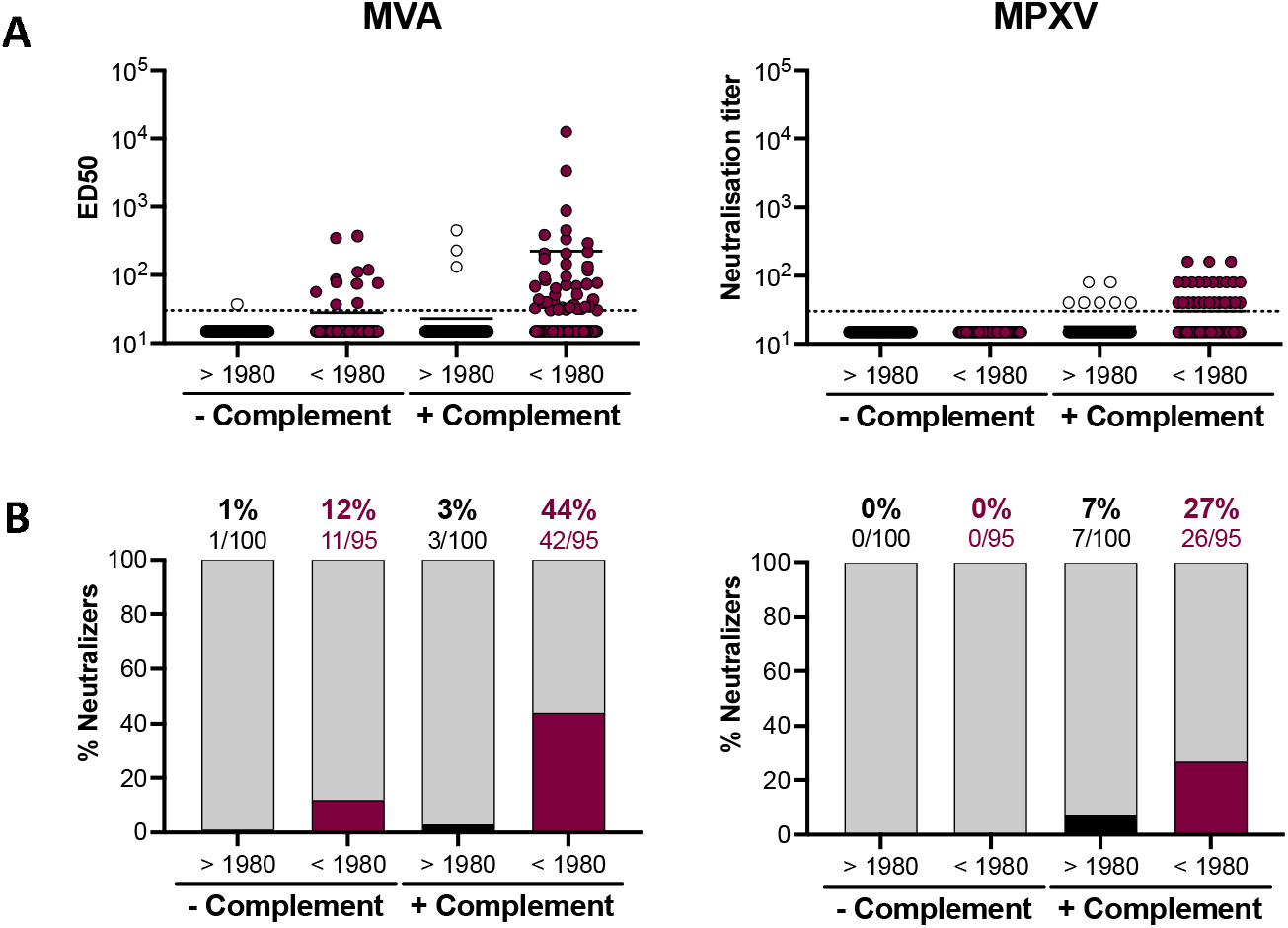
Basal levels of neutralizing antibodies in unvaccinated and uninfected individuals. (A) Seroneutralization of MVA-GFP (left panel) and MPXV (right panel) by sera from unvaccinated/uninfected individuals in the absence and the presence of 10% guinea pig complement as a source of complement. Youngers and older individuals were distinguished using 1980 as a cut-off year. The anti-MVA neutralizing activity was expressed as the median effective dose (ED50) corresponding to the dilution of plasma reducing MVA-GFP infection by 50%. The anti-MPXV neutralizing activity was expressed as neutralizing titer corresponding to the highest dilution factor in which neutralization was observed. The dotted lines represent the limit of detection (LOD). (B) The proportion of neutralizers was estimated as the percentage of individuals exhibiting a neutralizing activity > LOD.

Altogether, these results indicate that uninfected and unvaccinated individuals born after 1980 did neither neutralize MVA nor MPXV in absence of complement. Based on the results obtained on younger individuals in the presence of complement, the specificity of the MVA and MPXV assays was estimated to be 97% and 93%, respectively. The complement enhanced the sensitivity of the two assays.

### Neutralizing antibodies induced by MPXV infection

We then measured the neutralizing activity of sera from MPXV-infected individuals collected at different time points after onset of symptoms (Fig. 2). We classified the individuals in the two same categories of age to analyze the influence of potential smallpox childhood vaccination on MPXV infection-elicited antibody titers. With the MVA assay performed without complement, we detected anti-MVA NAbs in approximatively 60 % of patients regardless of the time after the onset of symptoms (mean ED50: 134). Higher levels of NAbs (mean ED50: 1,522) were detected in older individuals, suggesting a potential boost effect of MPXV infection on memory B cell populations previously generated by smallpox vaccination. Anti-MVA neutralizing titers increased over time. In the presence of complement, up to 83% of the younger individuals and 100% of the older individuals displayed NAbs 2-12 weeks after the onset of symptoms, with higher neutralization titers in older (mean ED50: 14,486) than in younger (mean ED50: 302) patients. With the MPXV assay, we did not detect NAbs in the absence of complement in the younger individuals. In the older individuals, 67% (4/6) displayed NAbs, 2 weeks after onset of symptoms. Addition of complement increased anti-MPXV activity of the sera, with respectively 65% and 100% of the younger and older individuals harboring NAbs 2 to 12 weeks after onset of symptoms (mean neutralization titers: 71 in younger and 600 in older individuals). With both MVA and MPXV assays, NAb titers were generally higher in older individuals. We then analyzed the evolution of NAb titers over time regardless of the age of the patients, by mixing the results obtained with the two age categories (Extended Data Fig. 4A and 4B). Both MVA- and MPXV-neutralizing titers increased over time. The sensitivity of the MVA and MPXV complement-based assays, assuming that all infected individuals have developed a neutralizing response when assessed ≥4 weeks after onset of symptoms, was estimated to be 94% and 82%, respectively. Neutralizing antibodies were detected in a majority (61%) of infected individuals as soon as one week after onset of symptoms.

**Fig. 2.**
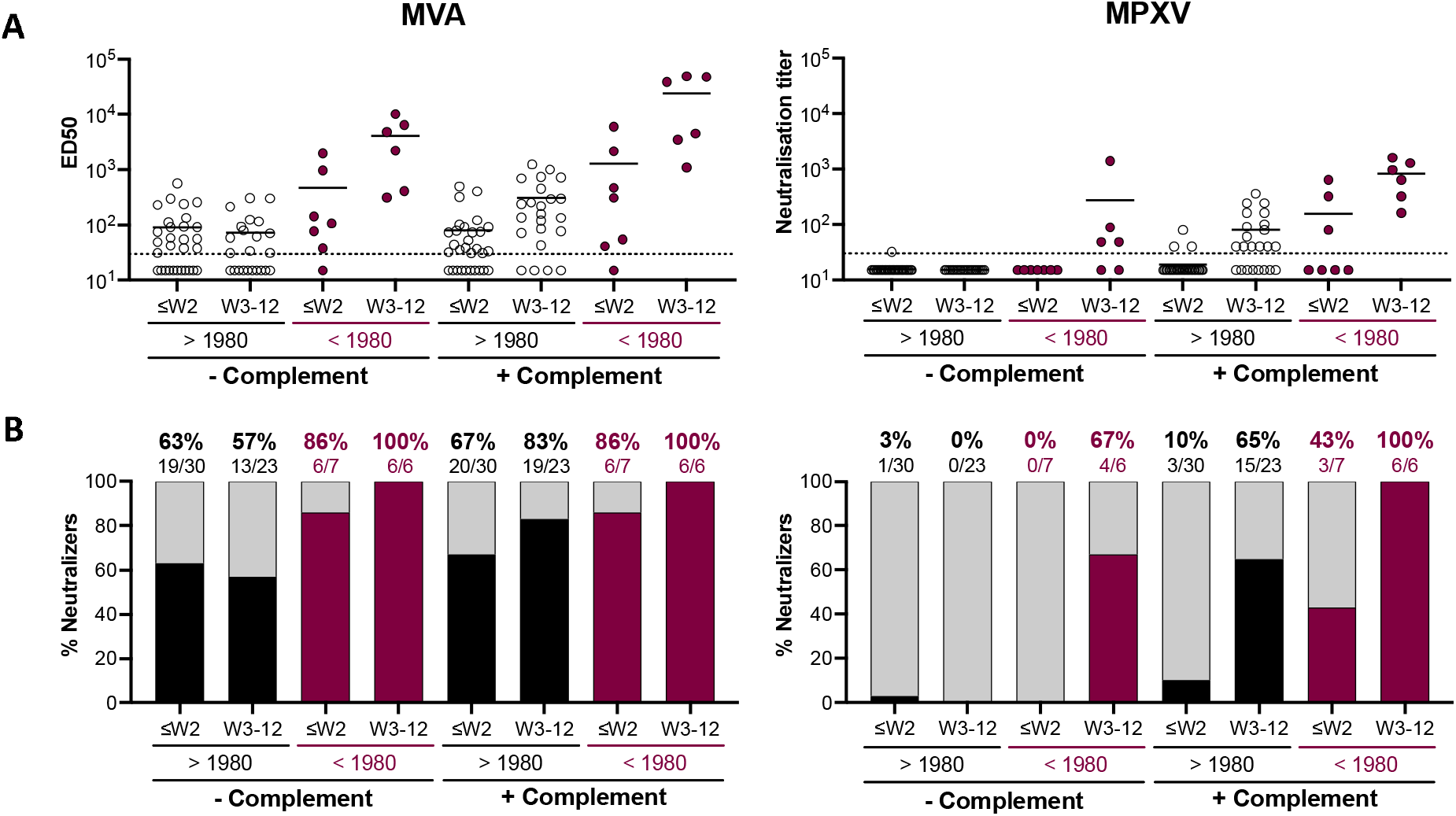
Neutralizing antibodies induced by MPXV infection. (A) Seroneutralization of MVA-GFP (left panel) and MPXV (right panel) by sera from MPXV-infected patients in the absence and presence of 10% guinea pig complement as a source of complement. Youngers and older individuals were distinguished using 1980 as a cut-off year. The anti-MVA neutralizing activity was expressed as the median effective dose (ED50) corresponding to the last dilution of plasma reducing MVA-GFP infection by 50%. The anti-MPXV neutralizing activity was expressed as neutralizing titer corresponding to the highest dilution factor in which neutralization was observed. In each category, individuals were separated according to the weeks (W) of sample collection (≤W2: samples collected during the 1^st^ or the 2^nd^ week of symptoms; W3-12: samples collected between the 3^rd^ and the 12^th^ week of symptoms). The dotted lines represent the limit of detection (LOD). (B) The proportion of neutralizers was estimated as the percentage of individuals exhibiting a neutralizing activity > LOD.

### Neutralizing antibodies elicited by IMVANEX vaccination

We next examined the ability of IMVANEX vaccination to elicit MVA-neutralizing antibodies (Fig. 3). The recent IMVANEX vaccination scheme in France consisted of two doses with an interval of 28 to 35 days in individuals born after 1980, or one single dose in individuals having received the historic smallpox vaccine. However, 4 older individuals (born before 1980) received two doses of IMVANEX and were also included in the analysis. With the MVA assay, we observed a progressive increase of anti-MVA NAbs after one and two vaccine doses. After 1 dose, the proportion of neutralizers was 17% and 77 % among younger and older participants, respectively. After 2 doses, the proportion reached respectively 71 % and 100 % of the younger and older individuals, highlighting the importance of the second dose. A single dose of MVA vaccine elicited similar levels of NAbs and a similar proportion of neutralizers in vaccinees that received the historic smallpox vaccine (mean ED50 of 209 in 77% of neutralizers) when compared to a two-dose regimen in the naïve individuals (mean ED50 of 136 in 71% of neutralizers). Addition of complement largely increased NAb titers, with 90 % of neutralizers detected among the young individuals (mean ED50: 470) and 100% among the older individuals (mean ED50: 1,890) 2 to 8 weeks after full vaccination. In younger individuals, anti-MVA neutralizing titers were 1.5-fold higher in vaccine recipients than in MPXV-infected patients. The same sera were tested in the MPXV-based assay. In the presence of complement, 52% of younger individuals having received two doses of IMVANEX and 40% of older individuals having received one dose of IMVANEX displayed detectable and similar levels of anti-MPXV Nabs (mean neutralization titers: 233 and 253, respectively). Anti-MPXV neutralizing titers were 3-fold higher in vaccinated individuals than in MPXV-infected patients.

**Fig. 3.**
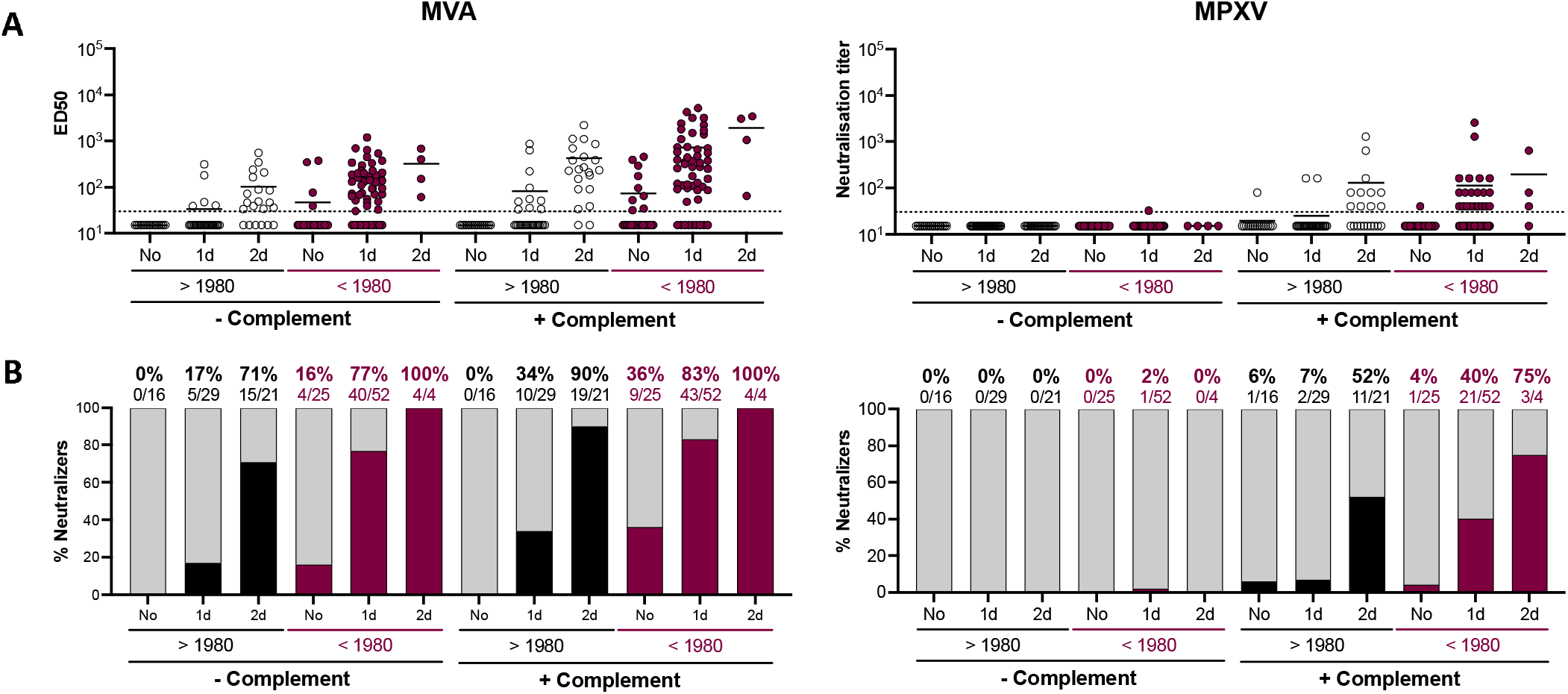
Neutralizing antibodies induced by IMVANEX vaccination. (A) Seroneutralization of MVA-GFP (left panel) and MPXV (right panel) by sera from IMVANEX vaccinees in the absence and presence of 10% guinea pig complement as a source of complement. Youngers and older individuals were distinguished using 1980 as a cut-off year. The anti-MVA neutralizing activity was expressed as the median effective dose (ED50) corresponding to the last dilution of plasma reducing MVA-GFP infection by 50%. The anti-MPXV neutralizing activity was expressed as neutralizing titer corresponding to the highest dilution factor in which neutralization was observed. In each category, In each category, individuals were separated according to their vaccination status at the time of sample collection (No : samples collected before vaccination; 1: after the 1st dose; 2: after the 2nd dose). The dotted lines represent the limit of detection (LOD). (B) The proportion of neutralizers was estimated as the percentage of individuals exhibiting a neutralizing activity > LOD.

These results indicate that IMVANEX elicited NAbs in most of vaccine recipients, with higher titers detected in older individuals. When combining the results obtained with the younger and older individuals in the two assays, we detected a proportion of 92% and 56% of vaccine recipients displaying anti-MVA and -MPXV NAbs, respectively. The sera neutralized more efficiently MVA than MPXV, reflecting the higher sensitivity of the MVA assay and/or a better anti-MVA immune response generated with an MVA-based vaccine.

### Neutralizing antibodies elicited by an experimental MVA-HIV vaccine

To further assess MVA immunogenicity, we measured NAb levels in sera from participants of a clinical trial (Fig. 4) that aimed at characterizing a candidate MVA vaccine carrying HIV antigens (MVA-HIV)^45^. Participants received two doses of the MVA-HIV vaccine candidate separated by 8 weeks. Sera were longitudinally collected before the first injection and 2 weeks after the first and second doses. In the two age categories, levels of anti-MVA NAbs increased after each injection and 94 to 98% of neutralizers were detected after 2 doses, regardless of the presence of complement. The proportion of neutralizers among MVA-HIV recipients was roughly similar to that of IMVANEX. In the younger individuals, anti-MVA neutralizing titers were about 3.6-fold higher 2 weeks after the second dose of MVA-HIV than 2 to 8 weeks after the second dose of IMVANEX. In the older individuals, the titers were similar after one dose of MVA-HIV or IMVANEX. When analyzed in the MPXV-based assay, NAb levels and the proportion of neutralizers were lower than those measured in the MVA-based assay. After two doses, 36% and 71% of younger and older individuals mildly neutralized MPXV, respectively.

**Fig. 4.**
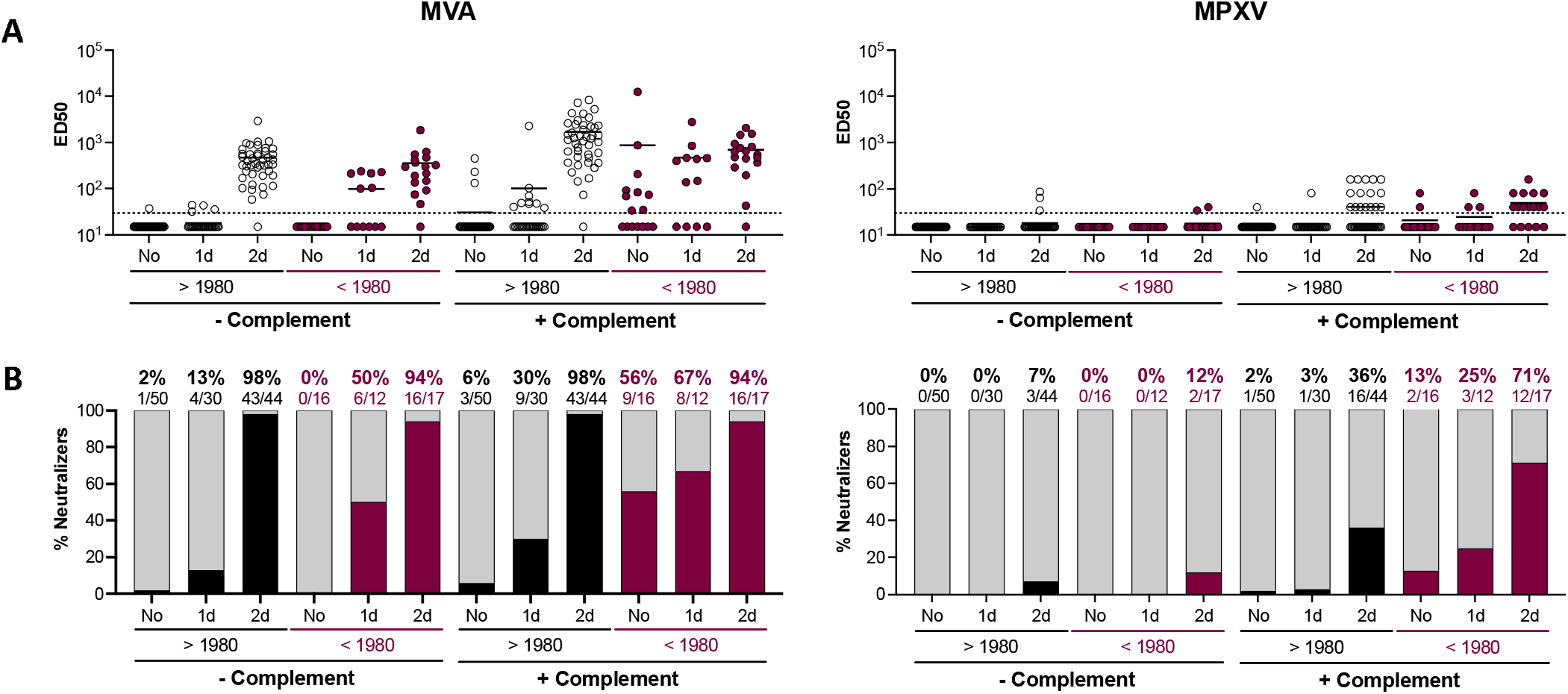
Neutralizing antibodies induced by an experimental MVA-HIV vaccine. (A) Seroneutralization of MVA-GFP (left panel) and MPXV (right panel) by sera from IMVANEX vaccinees in the absence and presence of 10% guinea pig complement as a source of complement. Youngers and older individuals were distinguished using 1980 as a cut-off year. The anti-MVA neutralizing activity was expressed as the median effective dose (ED50) corresponding to the last dilution of plasma reducing MVA-GFP infection by 50%. The anti-MPXV neutralizing activity was expressed as neutralizing titer corresponding to the highest dilution factor in which neutralization was observed. In each category, In each category, individuals were separated according to their vaccination status at the time of sample collection (No : samples collected before vaccination; 1: after the 1st dose; 2: after the 2nd dose). The dotted lines represent the limit of detection (LOD). (B) The proportion of neutralizers was estimated as the percentage of individuals exhibiting a neutralizing activity > LOD.

Therefore, despite potential differences in the MVA strain, preparation methods and injected doses, both IMVANEX and an experimental MVA-HIV vaccine triggered a similar and strong humoral response, mostly detectable in the presence of complement. NAbs titers were lower against MPXV than MVA.

### Correlation between MVA- and MPXV-based neutralization assays

We examined the links that may exist between anti-MVA and anti-MPXV NAb titers (Extended data Fig. 4C). There was a significant correlation between the two assays in infected individuals. The correlation was less marked but also significant in vaccine recipients. These data highlight the interest to use the more sensitive MVA-GFP system to monitor levels of poxvirus-specific antibodies.

## DISCUSSION

The efficacy of currently available third generation MVA-based vaccines against mpox and the nature of the humoral response generated after MPXV infection remain poorly characterized. We established two cell-based assays to measure the levels of NAbs targeting MVA or MPXV in ancient smallpox-vaccinated individuals, MPXV-infected patients, and IMVANEX or MVA-HIV vaccine recipients. We show that about 44% and 27% of French individuals born before 1980 carry detectable anti-MVA and anti-MPXV antibodies, respectively, confirming the existence of long-term cross-neutralizing antibodies elicited by childhood vaccination. Whether this residual neutralization protects against mpox disease could be assessed by comparing the clinical features of younger and older infected patients, if their historical vaccination status is known. Anti-MVA NAbs were detected in 94% of MPXV-infected patients up to 12 weeks after onset of illness (83% and 100% of younger and older patients, respectively), 92% of IMVANEX recipients (90% and 100% of younger and older individuals, respectively) and 97% of MVA-HIV recipients (98% and 94% of younger and older individuals, respectively) when assays were performed in the presence of complement. Addition of complement also enhanced anti-MPXV NAbs levels and the proportion of NAb-positive individuals, which reached 82% of MPXV-infected patients (65% and 100% of younger and older patients, respectively), 56% of IMVANEX recipients (52% and 75% of younger and older patients, respectively), and 46% of MVA-HIV recipients (36% and 71% of younger and older individuals, respectively). The differences in neutralization titers observed in younger and older individuals likely reflect the impact of childhood smallpox vaccination. Anti-MVA and MPXV NAb titers reached higher levels in older than in younger individuals, both after MPXV infection or first and second IMVANEX injection, strongly suggesting a reactivation of a long-term B cell memory response^21^. It is also possible that a hybrid immunity^52,53^, generated by a combination of vaccination and infection, triggers an efficient humoral response. The highest anti-MVA and anti-MPXV NAb titers were measured in ancient smallpox-vaccinated MPXV-infected patients, highlighting this potential optimal cross-protection mediated by hybrid immunity. The relatively low antibody levels with poor neutralizing capacities recently reported in Dutch cohorts of infected or vaccinated individuals might be due to the absence of use of complement in neutralizing assays^33^. Our work highlights the potential role of complement in improving the sensitivity of the assays and in more accurately reflecting the anti-poxvirus humoral response in infected or vaccinated individuals.

The mechanisms underlying anti-MPXV activity of the complement remain to be elucidated. The complement enhances VACV neutralization by monoclonal antibodies or sera from VACV-infected mice or vaccinated individuals^35–42^. Two infectious forms of viral particles are generated during the life cycle of poxviruses. Mature virions (MV) accumulate in the interior of infected cells and are released upon cell lysis while enveloped virions (EV) are wrapped MV secreted in the extracellular medium by exocytosis^54^. Due to their different morphogenesis, MV and EV express distinct sets of proteins at their surface. Twenty viral proteins decorate the surface of MV, including the entry-fusion complex composed of 11 viral proteins (A21, A28, G3, H2, O3, A16, F9, G9, J5, L1, L5)^55^, whereas EV contain 6 viral proteins (including A33, A34, and B5)^56^ anchored in the outer membrane. The fusogenic properties of MV make this form responsible for viral entry into target cells and more sensitive to neutralizing antibodies than EV. Immunodominant viral proteins are found in either MV (H3, A27, D8, L1) and EV (A33, B5)^49,57–62^. A recent study demonstrated that MPXV infection elicited strong antibody and B cell responses against A35 and H3 antigens^63^, which exhibit 95% and 93% homology with A33 and H3 VACV antigens, respectively. Neutralization of MV is easier to achieve than EV. The mechanisms of complement-mediated enhancement of neutralization vary between viral proteins. Anti-A33 antibodies induce complement-dependent virolysis of EV and subsequent release of MV^41^. Anti-B5 antibodies successively bind C1q and C3 in an isotype-dependent manner, leading to virion opsonization and sterically preventing EV attachment to target cells^35,42^. Binding of anti-B5 antibodies to VACV-infected cells leads to complement-dependent cytotoxicity (CDC)^35,42^. In a mouse model, an optimal protection against VACV challenge was mediated by a mixture of monoclonal antibodies that target 4 viral antigens^49^. Antibodies targeting A33, L1, A27 and H3 proteins exhibited the broadest cross-neutralizing activity against various OPXV including VACV, Cowpox virus (CPXV), and MPXV^49^. Future studies are warranted to characterize the role of the complement pathway during MPXV replication and its neutralization by polyclonal and monoclonal antibodies.

The titers of NAbs and the proportion of reactive individuals were globally higher with the MVA assay than with the MPXV assay. The greater sensitivity of the MVA assay is probably in part due to technical differences with the MPXV assay. MVA is a single-cycle virus in most mammalian cells, including the Vero E6 cells used here, and is thus likely more sensitive to neutralization than the fully replicative MPXV isolate. A direct cell-to-cell viral transfer^51^, visualized by clusters of MPXV infected cells with heterogenous sizes in our cultures, may also introduce a variability in the readout and may decrease NAb efficiency. It would have been of interest to measure the overall levels of anti-poxvirus antibodies in the different samples analyzed here. The lack of a validated commercially available ELISA assay at the initiation of this study, and the limited volumes of the sera precluded this analysis. It will be worth comparing our assays with ELISA tests or other serological assays in various groups of vaccinated or infected individuals.

The different sensitivities of the MVA and MPXV assays may also reflect antigenic diversity among the *Orthopoxvirus* genus. Anti-MPXV NAb titers were about two-fold higher in MPXV infected individuals than in MVA-based vaccine recipients, suggesting that the repertoire of cross-reactive antibodies generated upon MPXV infection may be different than after vaccination. However, anti-MVA NAb titers were of similar level in infected and vaccinated individuals. The MVA-based assay thus represents a sensitive and convenient tool to assess the anti-pox neutralizing response in immunized persons.

MVA-based vaccination (IMVANEX or JYNNEOS) is currently recommended as pre-exposure prophylaxis in high-risk populations or as post-exposure prophylaxis, even if breakthrough infections were observed after post-exposure vaccination^64^. Mpox incidence was 7.4- and 9.6-times lower in individuals having received on one or two doses of JYNEOS, respectively^65^, suggesting an impact of vaccination on mpox outbreak decline. The clinical forms of mpox were also milder in recently vaccinated than in unvaccinated patients^66^. Associating NAbs levels to clinical vaccine efficacy will help defining correlates of protection against MPXV infection or disease severity. MVA is also currently used as a candidate vaccine platform against several viruses, bacteria, and parasites^23^. The utilization of data from clinical trials with these candidates is of interest to define optimal vaccinal schemes to induce cross-neutralizing MPXV antibodies. For instance, we show that in the VRI01 trial, participants that received 2 doses of an MVA-based HIV vaccine candidate (MVA-HIV) elicited somewhat higher levels of anti-MVA NAbs than after IMVANEX administration. The differences might be due to various vaccine formulation, preparation, or administrated doses. An anti-OPXV cross-neutralizing response was also reported with other MVA-based vaccine candidates^33,67^.

Our study has several limitations. We did not have access to the historical smallpox vaccination status of the older individuals. However, the vaccination was obligatory in France until 1979 and it is likely that most of the participants were vaccinated. We did not measure the overall levels of anti-OPXV antibodies in the samples but focused on their neutralizing activity. We did not analyze the correlations that may exist between disease severity, HIV status and NAb levels, even if a preliminary survey did not reveal major differences between individuals, apart from the age effect reported here. It will be also of interest analyzing the long-term duration of the humoral response in the different categories of vaccinated or infected individuals.

In summary, we have analyzed the neutralizing humoral response elicited by MVA vaccination or MPXV infection. The sensitive neutralization assays that we implemented may help defining correlates of protection against infection or disease severity. The assays can be used for epidemiological surveys, assessment of the duration of protection conferred by previous infection or by authorized and candidate vaccines, and for analysis of any immunotherapeutic intervention.

## METHODS

### Cohort of healthy donors (CORSER-4)

A prospective, monocentric, longitudinal, interventional cohort clinical study (CORSER-4) is conducted since March 2020, with the objective to study the humoral responses after SARS-CoV-2 infection or vaccination in healthy individuals. This study was approved by the ethical committee “Comité de Protection des Personnes Ile de France III (CPP File N°Am8448-6-3765), on February 19, 2020. The cohort is registered with ClinicalTrials.gov (NCT04325646). At enrollment, written informed consent was collected from each individual, authorizing the analysis of seroneutralization against different viral species. We randomly selected 88 sera from 22- to 69-years old participants (Table 1). Smallpox vaccination was interrupted in 1980, the date of official smallpox eradication. Thus, we used 1980 as a birth year cut-off to distinguish “younger” and “older” healthy donors: 39% (34/88) were born after 1980 and were not vaccinated against smallpox (“younger”), whereas 61% (54/88) were born before 1980 (“older”) and probably received a historic smallpox vaccine.

### Hôpital Pitié-Salpêtrière cohort of MPXV-infected patients

In May 2022, a clinical study enrolling 70 individuals infected with MPXV was initiated^43,44^. Persons with PCR-confirmed mpox were invited to contribute to the case series by their health care provider. Written informed consent was obtained in accordance with local standards and maintained in the participants’ clinical file^43,44^. We obtained 57 sera or plasma collected from 39 patients. All patients were male. Eighteen (7/39) and 82% (32/39) were born before or after 1980, respectively. Samples were collected from 2 to 80 days after onset of symptoms, with a majority collected during the first two weeks of symptoms. 58% (33/57) were positive for MPXV by PCR and were collected from 2 to 25 days after onset of symptoms (median DOS: 5 days). Most of these samples were collected during the 1^st^ and 2^nd^ weeks of illness. 42% (24/57) were PCR-negative and were collected at a median DOS of 18 days. Three samples were collected later, at 47, 56 and 80 days post-symptoms.

### Hôpital Henri Mondor cohort of MPXV-infected patients

In July 2022, a prospective, monocentric cohort clinical study enrolling 9 individuals infected with MPXV was initiated. The study protocol was approved by the regional investigational review board (CPP Ile-de-France VII and IX) with approval reference 10–023. At enrollment, written informed consent was collected from each patient. For the present study, we obtained 9 sera from 9 patients (Table 2). All patients were male, 22% (2/9) and 78% (7/9) were born before and after 1980, respectively. Samples were collected from 18 to 33 days after the onset of symptoms and were negative for MPXV by PCR. Most of the samples (8/9) were collected at least 4 weeks after onset of symptoms.

### Orléans cohort of IMVANEX vaccinees

In April 2022, a prospective, monocentric, longitudinal, interventional cohort study enrolling up to 400 individuals was launched to explore humoral responses induced by either natural infection, vaccines, or therapeutic monoclonal antibodies. This cohort, firstly dedicated to COVID-19, was extended to various infectious diseases, and included persons vaccinated with IMVANEX. This study was approved by the “Comité de Protection des Personnes Est II” ethical committee under the reference 2022-A00177-36 (NCT NCT05315583). At enrollment, written informed consent was collected from each patient. Forty-two patients were included in this study: 40% (17/42) were born after 1980 and were not previously vaccinated against smallpox. They received two doses of IMVANEX vaccine with ≈30 days between each dose. Longitudinal samples were collected before vaccination, about 30 days after the 1^st^ dose and 35 days after the 2^nd^ dose (Table 3). Twenty nine percent (5/17) were treated for HIV and 59 % (10/17) were under antiretroviral pre-exposure prophylaxis (PrEP). Sixty percent (25/42) of the patients were born after 1980 and probably vaccinated against smallpox during childhood. The majority (22/25) received a single dose of IMVANEX whereas 12% (3/25) received a second dose. Sera were collected 34 days and 23 days after first and second doses, respectively. Sixty percent (15/25) were treated for HIV and 40 % (10/25) were under anti-retroviral PrEP.

### Hôpital Henri Mondor cohort of IMVANEX vaccinees

In July 2022, a prospective, monocentric, longitudinal cohort clinical study enrolling 43 individuals vaccinated with a third generation MVA-based vaccine (IMVANEX) was initiated. This study was approved by the appropriate ethical committee under the reference IDRCB: 2018-A01610-55. At enrollment, written informed consent was collected from each patient. For the present study, 43 patients were included: 35% (15/43) were born after 1980 and not previously vaccinated against smallpox. They received two doses of IMVANEX within 28 days of interval. Sera were collected 28 days after the 1^st^ dose and 15 days after the 2^nd^ dose (Table 3). Twenty-seven percent (4/15) were treated for HIV and 47% (7/15) were under pre-exposure prophylaxis (PrEP). Sixty five percent of the patients (28/43) were born before 1980 and probably vaccinated against smallpox during childhood. All received a single dose of IMVANEX and 3% (1/29) received two doses. Sera were collected 15 days after each dose. Among the older patients, 36% (10/28) were treated for HIV and 32% (9/28) were under pre-exposure prophylaxis (PrEP).

### Vaccine Research Institute cohort of MVA-HIV recipients

In 2014-2015, a clinical trial was performed in 92 individuals to evaluate the effectiveness of MVA-HIV as a vaccine candidate. The protocol was approved by an ethics committee (Comité de Protection des Personnes Ile-de-France V, Paris, France) and the competent French health authority (Agence Nationale de Sécurité du Médicament et des Produits de Santé) and was conducted in accordance with the declaration of Helsinki. All volunteers provided written and signed informed consent for the trial. The trial was registered with ClinicalTrials.gov (NCT02038842) and EudraCT (2012-002456-17)^45^. At enrollment, written informed consent was collected and each participant was exposed to two doses of MVA-HIV within 8 weeks of interval. Sera were collected before the initiation of the trial, 2 weeks after the 1^st^ dose, and 2 weeks after the 2^nd^ dose. We randomly selected 66 participants. Seventy-six percent (50/66) were born after 1980 and 24% (16/66) were older and probably vaccinated against smallpox during their childhood. A total of 169 sera was collected: 39% (66/169) before initiation of vaccination, 25% (42/169) after the first dose and 36% (61/169) after the second dose (Table 4).

### Cells and viruses

Vero E6 and U2OS cells^46^ were grown in Dulbecco’s Modified Eagle Medium (DMEM, Gibco) supplemented with 10% fetal bovine serum (FBS, Gibco), 100 U/mL penicillin, and 100 μg/mL streptomycin (Gibco). All cells were maintained at 37°C and 5% CO_2_ for culture. Cells were routinely tested negative for mycoplasma. A Modified Vaccinia virus Ankara carrying a GFP reporter gene (MVA-GFP)^47^ was provided by ANRS-MIE.

A MPXV strain (MPXV/2022/FR/CMIP) was isolated from a pustular lesion of a 36-year-old French man who consulted at the Medical Center of Institut Pasteur (CMIP), in June 2022. The clinical specimen was inoculated on Vero E6 cells, whose supernatant was harvested after 3 days and tested positive for the presence of MPXV by PCR^48^.

### MVA neutralization assay

Vero E6 cells were plated at 2□× □10^4^ cells per well in a μClear 96-well plate (Greiner Bio-One). Indicated concentrations of MVA-GFP were mixed (ratio 1:1) with serial dilutions (from 1/30 to 1/30,000) of previously heat-inactivated (30 min at 56°C) plasma or serum in the presence or absence of 10% guinea pig serum as a source of complement (GPC; Rockland). After incubation for 2 h at 37°C, the mixture was added onto Vero E6 cell monolayers. Twenty hours later, cells were fixed for 30 min at room temperature (RT) with 4% paraformaldehyde (PFA, Electron Microscopy Sciences), washed and stained with Hoechst (1:1,000 dilution; Invitrogen). Images were acquired with an Opera Phenix high-content confocal microscope (Perkin Elmer). The GFP area and the number of nuclei were quantified using the Harmony software (Perkin Elmer). The percentage of neutralization was calculated using the GFP area as the value with the following formula: 100□× □ (1□− □ (value with serum□− □value in “noninfected”)/(value in “no serum” □− □value in “noninfected”)). Neutralizing activity of each plasma or serum was expressed as the ED50 (effective dose inhibiting 50% of infection). ED50 were calculated using a reconstructed curve with the percentage of neutralization at the different serum concentrations.

### MPXV neutralization assay

U2OS cells were plated at 2□× □10^4^ cells per well in a μClear 96-well plate (Greiner Bio-One). Indicated concentrations of MPXV were mixed (ratio 1:1) in a BSL-3 facility with serial dilutions of previously heat-inactivated (30 min at 56°C) plasma or serum in the presence or absence of 10% GPC. After incubation for 2 h at 37°C, the mixture was added onto U2OS cells monolayers. Forty-eight hours later, cells were fixed for 30 min at RT with 4% paraformaldehyde, washed and immunostained for MPXV antigens with rabbit polyclonal anti-VACV antibodies (PA1-7258, Invitrogen), and an Alexa Fluor 488-coupled goat anti-rabbit antibody (Invitrogen). Nuclei were stained with Hoechst. Images were acquired with an Opera Phenix high-content confocal microscope (PerkinElmer). The MPXV^+^ area and the number of nuclei were quantified using the Harmony software (PerkinElmer). The neutralization titer was determined as the highest plasma or serum dilution in which the MPXV^+^ area was inferior to that of the “No serum” condition.

### Reproducibility of the MVA and MPXV neutralization assays

Each serum or plasma sample was tested at 6 dilutions in 2-4 independent experiments. Data are presented as the mean ED50 or neutralization titers obtained in the independent experiments. The intra-sample variability was below 5% in >95% of the samples. Samples were retested in case of discrepant results.

## Statistical analysis

Calculations were performed using Excel 365 (Microsoft). Figures and statistical analyses were conducted using GraphPad Prism 9. Statistical significance between different groups was calculated using the tests indicated in each figure legend. No statistical methods were used to predetermine sample size.

### Biosafety

All experiments with infectious MPXV were conducted under strict BSL3 conditions. Manipulations involving inactivated or non-inactivated MPXV were performed according to the French regulations on dual use pathogens.

## Data Availability

All data produced in the present work are contained in the manuscript.

## ACKNOWLEDGEMENTS

The authors thank the individuals and patients who participated to this study, the members of the Virus and Immunity Unit (Institut Pasteur) and other teams for discussion and help, Bruno Hoen and Pierre Buffet (Institut Pasteur) for their support, Quentin Grassin, Damien Hoinard and Maxence Feher (Institut Pasteur) for technical assistance and biosafety support, Nathalie Aulner and the staff at the UtechS Photonic BioImaging (UPBI) core facility (Institut Pasteur), a member of the France BioImaging network, for image acquisition and analysis, Emmanuel Roux, Marie-Charlotte Hallouin-Bernard (Institut Pasteur), Fabienne Peira, Vanessa Legros, Aurelie Theillay, Sandra Pallay, Daniela Pires Roteia (CHU Orléans), Agathe Nouchi, Vincent Berot, Yara Wakim, Cécile Brin, Ariane Gavaud, Yanis Tamzali, Alexandre Bleibtreu (Hôpital Pitié-Salpêtrière, APHP) and Véronique Rieux (ANRS-MIE) for their help with the cohorts and William Henry Bolland for critical reading of the manuscript.

## AUTHOR CONTRIBUTIONS

<u>Conceptualization:</u> M.H., F.G-B., T.B., F.P., D.P., O.S., L.H., V.P., T.P., A-G.M., J-D.L., C.B., Y.L., J-C.M., O.S.

<u>Methodology:</u> M.H., F.G-B., F.P., D.P., T.B., O.S.

<u>Investigation:</u> M.H., F.G-B., F.P., D.P., T.B., O.S.

<u>Data collection and analysis:</u> M.H., F.G-B., F.P., D.P., T.B.

<u>Cohorts design:</u> A.W., S.B., S.M., R.P., G.M., H.D., S.G., J-L.L-Z., W.V., F.T., S.F.-P., M.D., H.L., L.A., M-N.U., L.H., V.P., T.P., A-G.M., J-D.L., Y.L.

<u>Resources:</u> J.V., A.W., S.B., S.M., R.P., G.M., H.D., S.G., J-L.L-Z., W.V., F.T., S.F.-P., M.D., H.L., L.A., M-N.U., L.H., V.P., T.P., A-G.M., J-D.L., C.B., Y.L., J-C.M.

<u>Manuscript writing and editing:</u> M.H., F.G-B., T.B., A.W., S.B., R.P., L.H., V.P., T.P., A-G.M., J-D.L., C.B., Y.L., J-C.M., O.S.

## FIGURE TITLES

**Extended Data Fig. 1**. Associations between time after onset of symptoms, day of mpox diagnosis and detection of MPXV genome in plasma samples from MPXV-infected patients

**Extended Data Fig. 2**. The MVA-GFP seroneutralization assay

**Extended Data Fig. 3**. The MPXV seroneutralization assay

**Extended Data Fig. 4**. Evolution and correlation of MPXV infection-elicited antibodies overtime

## FIGURE LEGENDS

**Extended Data Fig. 1. Associations between time after onset of symptoms, day of mpox diagnosis and detection of MPXV genome in plasma samples from MPXV-infected patients**

(A) Detection of MPXV genome by PCR in the plasma of MPXV-infected patients at the indicated days after onset of symptoms (DOS) (n=66)

(B) Detection of MPXV in the plasma of infected patients at the indicated weeks after onset of symptoms.

(C) Correlation between time after onset of symptoms and time after mpox diagnosis. The red oval shows the delay between symptoms and diagnosis.

**Extended Data Fig. 2. The MVA-GFP seroneutralization assay**

(A) A Modified Vaccinia virus Ankara carrying a GFP reporter gene (MVA-GFP) was mixed to serial dilutions of sera from unvaccinated/uninfected individuals, MPXV-infected patients, IMVANEX vaccinees or MVA-HIV recipients in the absence or the presence of 10% guinea pig complement. The mixture was then incubated for 30min at room temperature (RT) or 2h at 37°C before addition of Vero E6 target cells. After 20h, cells become GFP^+^ upon infection and the GFP^+^ infected cells were quantified by high-content confocal microscopy. The percentage of neutralization was calculated as mentioned in the Material and Methods section.

(B) Vero E6 cells were exposed to several doses of MVA-GFP. After 20h, the GFP^+^ area of infected cells was quantified by fluorescence microscopy. Left: representative images of the MVA-GFP-infected cells according to the dose of viral inoculum. The numbers indicate the dilution factors of the viral suspension. Right: standard curve and linearity of the GFP signal following MVA-GFP infection.

(C) Examples of the neutralization of MVA-GFP by 4-fold serial dilutions of two plasma from the same MPXV-infected patient at 2 days (non-neutralizing plasma) and 56 days (neutralizing plasma) after the onset of symptoms. Left: examples of micrographs and the white numbers show the percentage of neutralization by considering the “No serum” and “Non infected” conditions as 0% and 100% neutralization, respectively. Right: representative graph of the neutralization profiles of the non-neutralizing serum (in red) and the neutralizing serum (in grey). The median effective dose (ED50) was calculated for each plasma as the dilution of plasma which is necessary to reduce MVA-GFP infection by 50%, using the “No serum” condition as the basis for maximal infection.

(D) Example of the neutralization of several MVA-GFP doses by a serum from an IMVANEX vaccinee in the absence (left panel) or the presence (right panel) of 10% guinea pig complement. Representative graphs of the neutralization profiles.

(E) Comparison of the neutralization of MVA-GFP (dilution factor: 2.10^4^) by a serum from an IMVANEX vaccinee in the presence or the absence of complement.

**Extended Data Fig. 3. The MPXV seroneutralization assay**

(A) Mpox virus (MPXV) was mixed to serial dilutions of sera from unvaccinated/uninfected individuals, MPXV-infected patients, IMVANEX vaccinees or MVA-HIV recipients in the absence or the presence of 10% guinea pig complement. The mixture was then incubated for 30min at room temperature (RT) or 2h at 37°C before addition of U2OS target cells. After 48h, the cells were fixed and MPXV infection was monitored by immunofluorescence using a rabbit polyclonal anti-VACV antibody. The percentage of neutralization was calculated as mentioned in the Material and Methods section.

(B) U2OS cells were exposed to several doses of MPXV. After 48h, cells were fixed and MPXV was stained using a cross-reactive polyclonal anti-VACV antibody. Left: representative images of the MPXV-infected cells according to the dose of viral inoculum. Right: standard curve and linearity of MPXV infection.

(C) Examples of the neutralization of MPXV by 2-fold serial dilutions of a non-neutralizing serum (unvaccinated and uninfected individual) and a neutralizing serum (MPXV-infected patient). The white symbols (“-” and “+”) show the the presence (“+”) or the absence (“-”) of a neutralizing activity by considering neutralization every GFP area below the GFP area of the “No serum” condition. The neutralizing titer (red arrow) was determined by selecting the highest dilution in which neutralization was observed and was expressed as the corresponding dilution factor.

**Extended Data Fig. 4. Evolution and correlation of anti-MVA and anti-MPXV NAbs**

(A) Seroneutralization of MVA-GFP (left panel) and MPXV (right panel) by sera from MPXV-infected patients in the absence and presence of 10% guinea pig complement as a source of complement. Youngers and older individuals were distinguished using 1980 as a cut-off year. The anti-MVA neutralizing activity was expressed as the median effective dose (ED50) corresponding to the last dilution of plasma reducing MVA-GFP infection by 50%. The anti-MPXV neutralizing activity was expressed as neutralizing titer corresponding to the highest dilution factor in which neutralization was observed. In each category, individuals were separated according to the weeks (W) of sample collection. The dotted lines represent the limit of detection (LOD).

(B) The proportion of neutralizers was estimated as the percentage of individuals exhibiting a neutralizing activity > LOD.

(C) Correlative analysis of the neutralizing activity of sera from MPXV-infected patients, IMVANEX vaccinees and MVA-HIV patients against MVA and MPXV.

