## Supplementary files for "Complement-dependent mpox virus-neutralizing antibodies in infected and vaccinated individuals"

### Slide 1
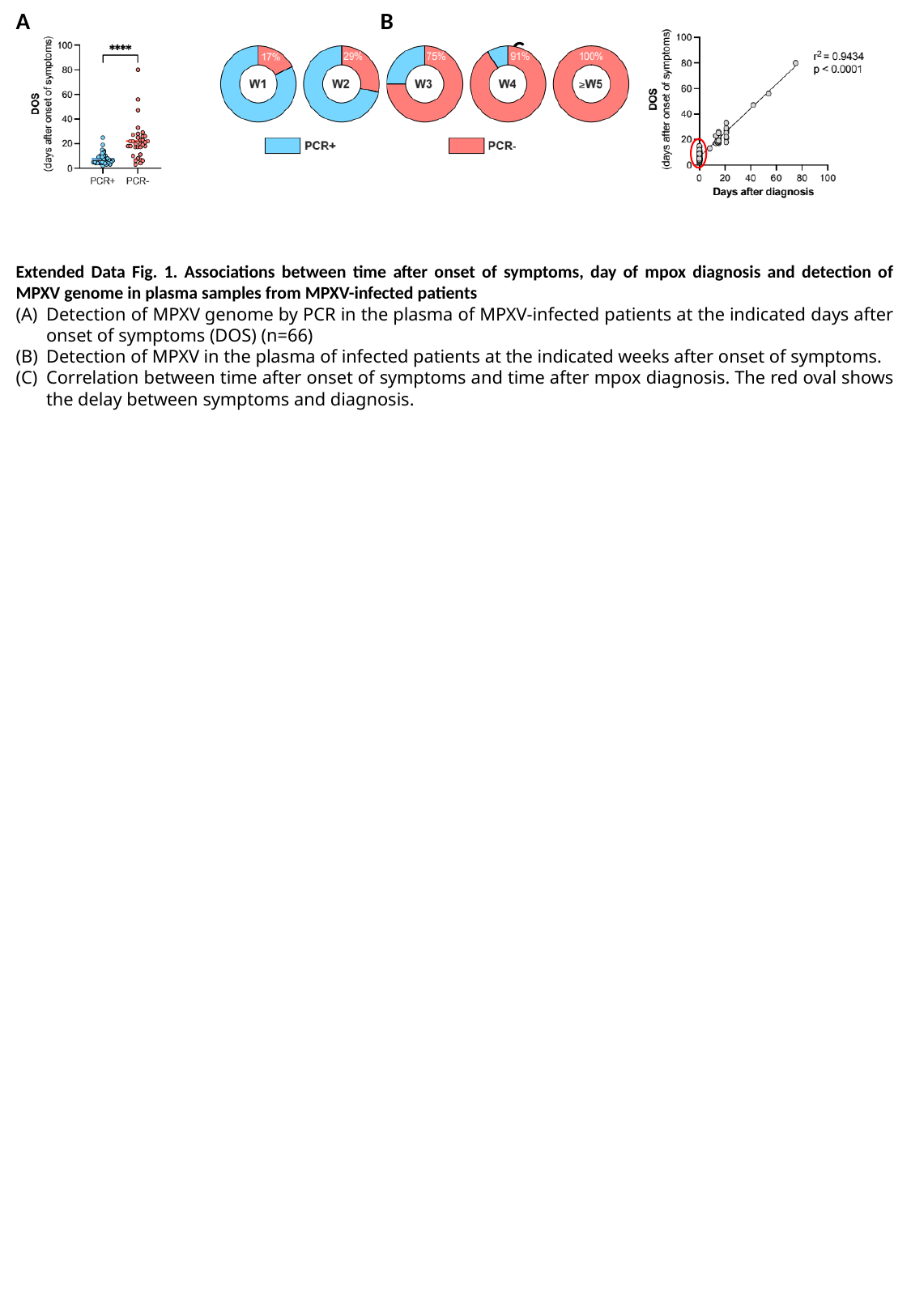

A			B							 C
Extended Data Fig. 1. Associations between time after onset of symptoms, day of mpox diagnosis and detection of MPXV genome in plasma samples from MPXV-infected patients
Detection of MPXV genome by PCR in the plasma of MPXV-infected patients at the indicated days after onset of symptoms (DOS) (n=66)
Detection of MPXV in the plasma of infected patients at the indicated weeks after onset of symptoms.
Correlation between time after onset of symptoms and time after mpox diagnosis. The red oval shows the delay between symptoms and diagnosis.

### Slide 2
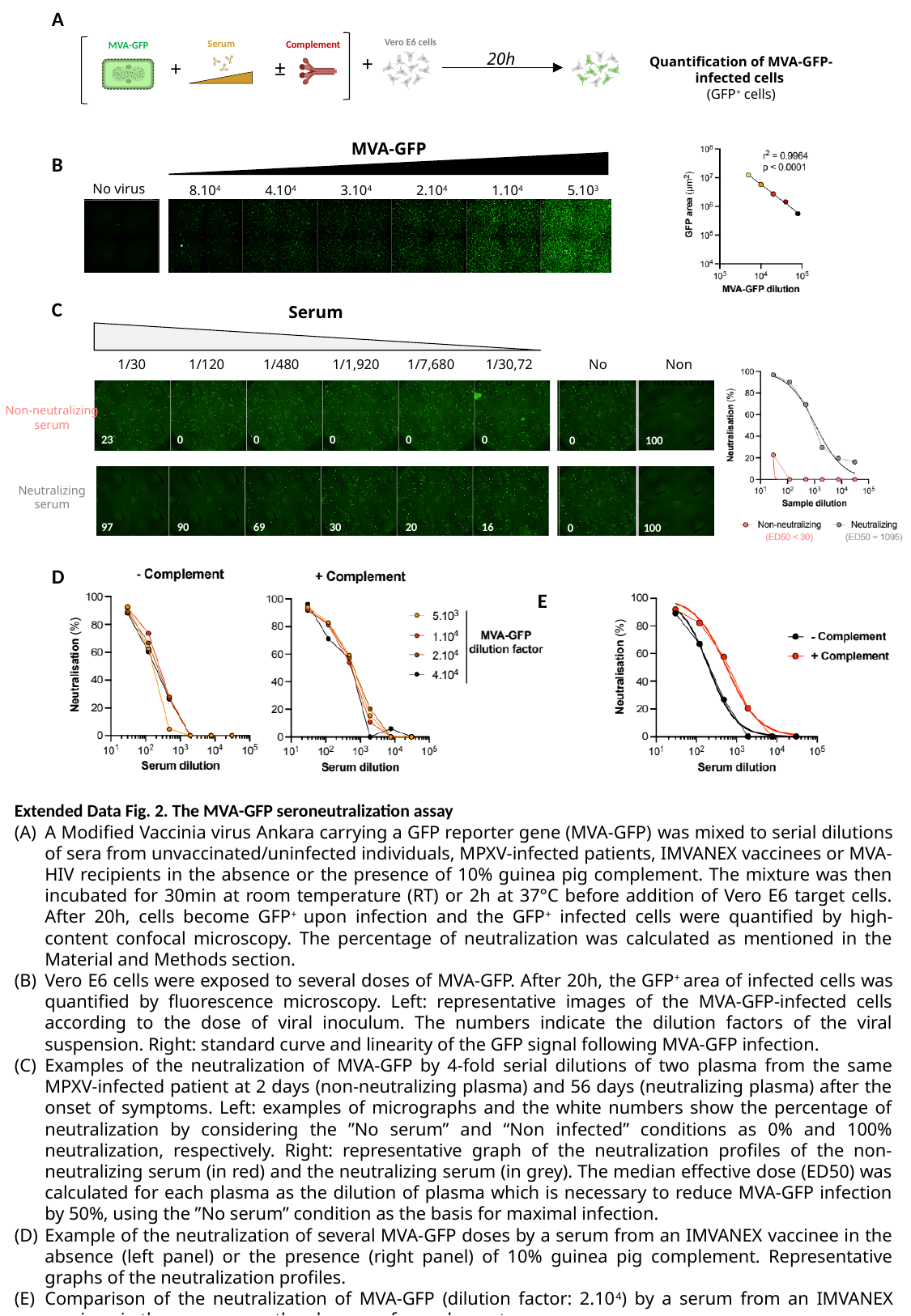

A
B
C
D									E
Vero E6 cells
Serum
Complement
MVA-GFP
20h
+
Quantification of MVA-GFP-infected cells
(GFP^+^ cells)
±
+
MVA-GFP
No virus
8.10^4^
4.10^4^
3.10^4^
2.10^4^
1.10^4^
5.10^3^
Serum
No serum
Non infected
1/30
1/120
1/480
1/1,920
1/7,680
1/30,720
Non-neutralizing
serum
23
0
0
0
0
0
0
100
Neutralizing
serum
97
90
69
30
20
16
0
100
Extended Data Fig. 2. The MVA-GFP seroneutralization assay
A Modified Vaccinia virus Ankara carrying a GFP reporter gene (MVA-GFP) was mixed to serial dilutions of sera from unvaccinated/uninfected individuals, MPXV-infected patients, IMVANEX vaccinees or MVA-HIV recipients in the absence or the presence of 10% guinea pig complement. The mixture was then incubated for 30min at room temperature (RT) or 2h at 37°C before addition of Vero E6 target cells. After 20h, cells become GFP^+^ upon infection and the GFP^+^ infected cells were quantified by high-content confocal microscopy. The percentage of neutralization was calculated as mentioned in the Material and Methods section.
Vero E6 cells were exposed to several doses of MVA-GFP. After 20h, the GFP^+^ area of infected cells was quantified by fluorescence microscopy. Left: representative images of the MVA-GFP-infected cells according to the dose of viral inoculum. The numbers indicate the dilution factors of the viral suspension. Right: standard curve and linearity of the GFP signal following MVA-GFP infection.
Examples of the neutralization of MVA-GFP by 4-fold serial dilutions of two plasma from the same MPXV-infected patient at 2 days (non-neutralizing plasma) and 56 days (neutralizing plasma) after the onset of symptoms. Left: examples of micrographs and the white numbers show the percentage of neutralization by considering the ”No serum” and “Non infected” conditions as 0% and 100% neutralization, respectively. Right: representative graph of the neutralization profiles of the non-neutralizing serum (in red) and the neutralizing serum (in grey). The median effective dose (ED50) was calculated for each plasma as the dilution of plasma which is necessary to reduce MVA-GFP infection by 50%, using the ”No serum” condition as the basis for maximal infection.
Example of the neutralization of several MVA-GFP doses by a serum from an IMVANEX vaccinee in the absence (left panel) or the presence (right panel) of 10% guinea pig complement. Representative graphs of the neutralization profiles.
Comparison of the neutralization of MVA-GFP (dilution factor: 2.10^4^) by a serum from an IMVANEX vaccinee in the presence or the absence of complement.

### Slide 3
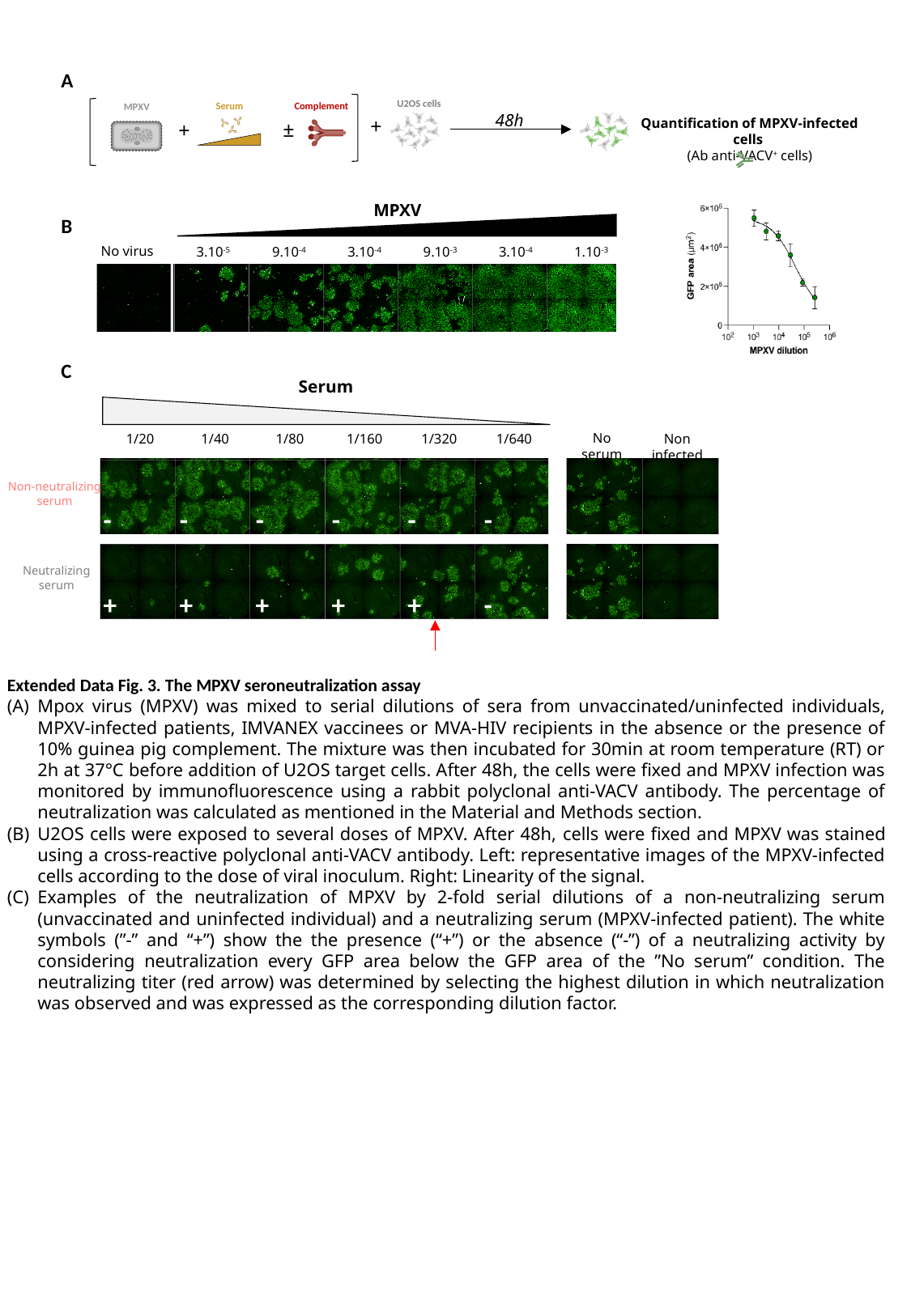

A
B
C
U2OS cells
Serum
Complement
MPXV
48h
+
Quantification of MPXV-infected cells
(Ab anti-VACV^+^ cells)
±
+
MPXV
No virus
3.10^-5^
9.10^-4^
3.10^-4^
9.10^-3^
3.10^-4^
1.10^-3^
Serum
No serum
Non infected
1/20
1/40
1/80
1/160
1/320
1/640
Non-neutralizing
serum
-
-
-
-
-
-
Neutralizing
serum
+
+
+
+
+
-
Extended Data Fig. 3. The MPXV seroneutralization assay
Mpox virus (MPXV) was mixed to serial dilutions of sera from unvaccinated/uninfected individuals, MPXV-infected patients, IMVANEX vaccinees or MVA-HIV recipients in the absence or the presence of 10% guinea pig complement. The mixture was then incubated for 30min at room temperature (RT) or 2h at 37°C before addition of U2OS target cells. After 48h, the cells were fixed and MPXV infection was monitored by immunofluorescence using a rabbit polyclonal anti-VACV antibody. The percentage of neutralization was calculated as mentioned in the Material and Methods section.
U2OS cells were exposed to several doses of MPXV. After 48h, cells were fixed and MPXV was stained using a cross-reactive polyclonal anti-VACV antibody. Left: representative images of the MPXV-infected cells according to the dose of viral inoculum. Right: Linearity of the signal.
Examples of the neutralization of MPXV by 2-fold serial dilutions of a non-neutralizing serum (unvaccinated and uninfected individual) and a neutralizing serum (MPXV-infected patient). The white symbols (”-” and “+”) show the the presence (“+”) or the absence (“-”) of a neutralizing activity by considering neutralization every GFP area below the GFP area of the ”No serum” condition. The neutralizing titer (red arrow) was determined by selecting the highest dilution in which neutralization was observed and was expressed as the corresponding dilution factor.

### Slide 4
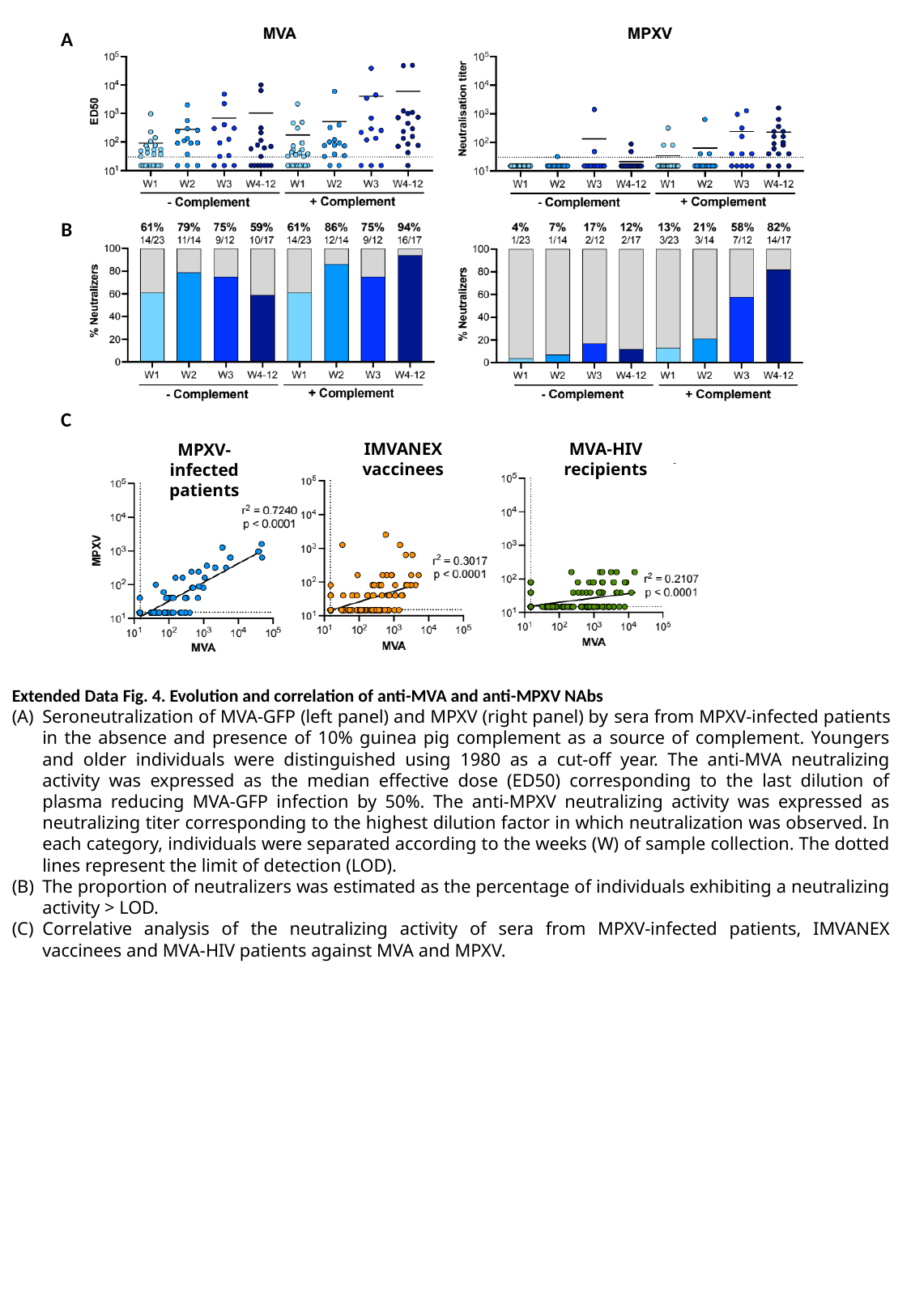

A
B
C
IMVANEX vaccinees
MVA-HIV recipients
MPXV-infected patients
Extended Data Fig. 4. Evolution and correlation of anti-MVA and anti-MPXV NAbs
Seroneutralization of MVA-GFP (left panel) and MPXV (right panel) by sera from MPXV-infected patients in the absence and presence of 10% guinea pig complement as a source of complement. Youngers and older individuals were distinguished using 1980 as a cut-off year. The anti-MVA neutralizing activity was expressed as the median effective dose (ED50) corresponding to the last dilution of plasma reducing MVA-GFP infection by 50%. The anti-MPXV neutralizing activity was expressed as neutralizing titer corresponding to the highest dilution factor in which neutralization was observed. In each category, individuals were separated according to the weeks (W) of sample collection. The dotted lines represent the limit of detection (LOD).
The proportion of neutralizers was estimated as the percentage of individuals exhibiting a neutralizing activity > LOD.
Correlative analysis of the neutralizing activity of sera from MPXV-infected patients, IMVANEX vaccinees and MVA-HIV patients against MVA and MPXV.
