## Supplementary material for "Complement-dependent mpox virus-neutralizing antibodies in infected and vaccinated individuals": Tables

| **Nb of patients** | 88 |
| --- | --- |
| **Sex** |  |
| Female | 53 (60%) |
| Male | 32 (37%) |
| Unknown | 3 (3%) |
| **Age** | 51 [22-69] |
| **Birth year** |  |
| **> 1980** | 34 (39%) |
| **< 1980** | 54 (61%) |

**Table 1. Characteristics of healthy donors**

|  | **Hôpital Pitié-Salpêtrière (n=39)** | |  | **Hôpital Henri Mondor (n=9)** | |
| --- | --- | --- | --- | --- | --- |
|  | **> 1980** | **< 1980** |  | **> 1980** | **< 1980** |
| **Nb of patients** | 32 (82%) | 7 (18%) |  | 7 (78%) | 2 (22%) |
| **Sex** |  |  |  |  |  |
| Female | 0 (0%) | 0 (0%) |  | 0 (0%) | 0 (0%) |
| Male | 32 (100%) | 7 (100%) |  | 7 (100%) | 2 (100%) |
| **Age** | 31 [21-41] | 51 [43-62] |  | 32 [24-40] | 57 [49-64] |
| **Days after onset of symptoms** | 10 [3-80] | 6 [2-56] |  | 27 [21-33] | 22 [18-26] |
| **Nb of samples** | 46 | 11 |  | 7 | 2 |
| Week 1 (W1) | 17 (37%) | 6 (55%) |  | 0 (0%) | 0 (0%) |
| Week 2 (W2) | 13 (28%) | 1 (9%) |  | 0 (0%) | 0 (0%) |
| Week 3 (W3) | 9 (20%) | 2 (18%) |  | 0 (0%) | 1 (50%) |
| Week 4 to 12 (W4-12) | 7 (15%) | 2 (18%) |  | 7 (100%) | 1 (50%) |

**Table 2. Characteristics of MPXV-infected patients**

|  | **CHR Orléans (n=42)** | |  | **Hôpital Henri Mondor (n=43)** | |
| --- | --- | --- | --- | --- | --- |
|  | **> 1980** | **< 1980** |  | **> 1980** | **< 1980** |
| **Nb of patients** | 17 (40%) | 25 (60%) |  | 15 (35%) | 28 (65%) |
| **Sex** |  |  |  |  |  |
| Female | 0 (0%) | 0 (0%) |  | 0% (0%) | 0% (0%) |
| Male | 17 (100%) | 25 (100%) |  | 15 (100%) | 28 (100%) |
| **Age** | 38 [21-41] | 55 [43-71] |  | 35 [22-41] | 54 [44-71] |
| **Status** |  |  |  |  |  |
| HIV positivity | 5 (29%) | 15 (60%) |  | 4 (27%) | 10 (36%) |
| HIV PrEP | 10 (59%) | 10 (40%) |  | 7 (47%) | 9 (32%) |
| **IMVANEX vaccine** |  |  |  |  |  |
| No dose | 16 (94%) | 25 (100%) |  | 0 (0%) | 0 (0%) |
| 1 dose | 17 (100%) | 24 (96%) |  | 12 (80%) | 28 (100%) |
| 2 doses | 13 (76%) | 3 (12%) |  | 8 (53%) | 1 (3%) |
| **Delay between doses** | 30 [28-42] | 34 [28-71] |  | 28 [28-28] | 28 [28-28] |
| **Time of serum collection** |  |  |  |  |  |
| After the 1^st^ dose | 30 [28-42] | 34 [28-71] |  | 28 [28-28] | 15 [15-28] |
| After the 2^nd^ dose | 35 [10-56] | 23 [22-40] |  | 15 [15-15] | 15 [15-15] |

**Table 3. Characteristics of IMVANEX vaccinees**

|  | **VRI (n=66)** | |
| --- | --- | --- |
|  | **> 1980** | **< 1980** |
| **Nb of participants** | 50 (76%) | 16 (24%) |
| **Sex** |  |  |
| Unknown | 50 (100%) | 16 (100%) |
| **Age** | N.A. | N.A. |
| **MVA-HIV vaccine** |  |  |
| No dose | 50 (100%) | 16 (100%) |
| 1 dose | 30 (60%) | 12 (75%) |
| 2 doses | 44 (88%) | 17 (100%) |
| **Delay between doses** | 8 weeks | 8 weeks |
| **Time of serum collection** |  |  |
| After the 1^st^ dose | 2 weeks | 2 weeks |
| After the 2^nd^ dose | 2 weeks | 2 weeks |

**Table 4. Characteristics of MVA-HIV recipients**
